# Protocol for a cluster randomised controlled trial to evaluate the effectiveness of digital health interventions in improving non-communicable disease management during the pandemic in rural Pakistan

**DOI:** 10.1101/2023.02.17.23286113

**Authors:** Xiaolin Wei, Nida Khan, Hammad Durrani, Naila Muzaffar, Victoria Haldane, John D. Walley, Kevin Thorpe, Erjia Ge, Shiliang Ge, Warren Dodd, James Wallace, Garry Aslanyan, Audrey Laporte, Muhammad Amir Khan

## Abstract

**Background:** The COVID-19 pandemic has revealed gaps in global health systems, especially in the low- and middle-income countries (LMICs). Evidence shows that patients with non-communicable diseases (NCDs) are at higher risk of contracting COVID-19 and suffering direct and indirect health consequences. Considering the future challenges such as environmental disasters and pandemics to the LMICs health systems, digital health interventions (DHI) are well poised to strengthen health care resilience. This study aims to implement and evaluate a comprehensive package of DHIs of integrated COVID-NCD care to manage NCDs in primary care facilities in rural Pakistan.

**Methods:** The study is designed as a pragmatic, parallel two-arm, multi-centre, cluster randomised controlled trial. We will randomise 30 primary care facilities in three districts of Punjab, where basic hypertension and diabetes diagnosis and treatment is provided, with a ratio of 1:1 between intervention and control. In each facility, we will recruit 50 patients who have uncontrolled hypertension. The intervention arm will receive training on an integrated COVID-NCD guideline, and will use a smartphone app-based telemedicine platform where patients can communicate with health providers and peer-supporters, along with a remote training and supervision system. Usual care will be provided in the control arm. Patients will be followed up for 10 months. Our primary indicator is systolic blood pressure measured at 10 months. A process evaluation guided by implementation science frameworks will be conducted to explore implementation questions. A cost-effectiveness evaluation will be conducted to inform future scale up in Pakistan and other LMICs.

**Discussion:** Our study is one of the first randomised controlled trials to evaluate the effectiveness of DHIs to manage NCDs to strengthen health system resilience in LMICs. We will also evaluate the implementation process and cost-effectiveness to inform future scale-up in similar resource constrained settings.

**Trial registration:** ClinicalTrials.gov Identifier - NCT05699369

## Introduction

Throughout the COVID-19 pandemic, delivering care for non-communicable diseases (NCDs) has been challenged, especially in low-and-middle-income countries (LMICs) with high burdens of NCDs such as Pakistan (1). Indeed, Pakistan ranks 7^th^ globally amongst countries with the highest number of persons living with diabetes and hypertension (2). People with diabetes and hypertension require ongoing care and contact with health providers. Yet in Pakistan, NCDs are inadequately managed in primary care, as evidenced by high rates of vascular complications and low blood pressure control rates (<30%) (3,4). The situation was made worse by the COVID-19 pandemic. Throughout COVID-19, accessing NCD care in Pakistan was made more difficult due to infection control and social distancing measures at health facilities (5). At the same time, people with these conditions are at greater risk of severe complications and poor health outcomes from COVID-19, prompting some to forgo care seeking to avoid infection (6,7).

The NCD Countdown 2030 specifically calls for focused scale-up of feasible and cost-effective interventions to ensure people living with NCDs can access the care they need, while supporting the dual goal of helping countries build back better and creating a sustainable foundation for a more ambitious global NCD agenda (8). Digital health interventions (DHIs), including telemedicine, mhealth, virtual care, and remote management have been used prior to the pandemic to provide feasible and effective integrated NCD care, especially in managing multimorbidity, delivering precision medicine, and increasing medication adherence (9). Building upon these prior efforts, digital health tools have emerged during the pandemic as an important strategy to improve health system resilience globally. A WHO rapid review conducted in May 2020 found that half of the 163 countries reviewed had disrupted their NCD services during the pandemic, and over half began to adopt DHIs such as telemedicine in their NCD services, with various adoptions in the US, China, and Iran (10,11). The implementation of DHIs has largely been a strategy to limit exposure to COVID-19 when seeking or providing care, reduce nosocomial and community transmission, and has emerged as acceptable to both health providers and patients (6,12). However, DHIs require robust information technology systems, privacy and security infrastructure, and importantly, reliable internet or mobile phone coverage and connectivity. Most primary care facilities in LMICs lack the capacity to develop qualified telemedicine platforms and ensure their ongoing availability (13). Given these requirements and constraints, limited functions of DHI, such as phone calls and text messages were reported as the most common form of DHIs, with fewer examples leveraging more advanced features such as smartphone app-based operations mostly applied in developed countries (14,15).

There are 185 million mobile connections in Pakistan among a total population of 225 million; and around half of these connections are smart phone users (16). In Pakistan, provincial governments have made efforts to support the development of DHIs. For instance, the Government of Sindh approved the Telemedicine and TeleHealth Bill 2021, which aimed to facilitate, enhance and improve access to healthcare through digital health platforms to ease the shortage of healthcare professionals (17). Pakistan has seen several DHI initiatives piloted by provincial governments and not-for-profit organisations. Telehealth clinics were established in some isolated and rural areas of Punjab, Sindh and Khyber-Pakhtunkhwa provinces to provide easy access to healthcare specialties such as dermatology, radiology and psychiatry (18–20). In a recent initiative, a tele-triage centre was established in Punjab with good success, focused on management and follow-up COVID-19 patients (21). However, the initiatives are all sporadic that there is no sustained design and plan to scale up DHIs at health system level in Pakistan. For longer-term success, DHIs must consider culture, norms and traditions in Pakistan to improve acceptability, and meet technical, procedural and operational needs (22).

Our previous studies in Pakistan demonstrated the feasibility and effectiveness of a community-based case management programme for patients with hypertension or diabetes (23,24). Based on this, we aim to design and implement a comprehensive package of DHI interventions to improve the management of patients with NCDs (hypertension and diabetes in this setting) in rural primary care facilities in Punjab Province, Pakistan during the pandemic. In addition, DHIs tested in this trial would strengthen the health system resilience and help to prepare other hazards, such as environment related disasters of flood as recently happened in Pakistan. We will also explore the implementation questions regarding feasibility and cost-effectiveness of this comprehensive package of DHIs for future scale-up in Pakistan and other LMICs to prepare all-purpose hazards.

## Material and Methods

This protocol is reported according to the SPIRIT guidelines (25).

### Study Design

We will conduct a parallel two-arm, multi-centre, cluster randomized controlled trial, using a 1:1 allocation ratio, to evaluate whether the DHI package is superior to usual care for blood pressure management among patients with NCDs. The study design is informed by the Medical Research Council framework (26) on complex interventions and implementation science frameworks with an embedded theory-based process evaluation to examine implementation questions regarding acceptability, reach, adoption, implementation and maintenance, as well as appropriateness and burden of technology use (27,28). In addition, we will conduct an incremental cost-effectiveness analysis to inform future scale-up. The trial protocol has obtained ethical approval from the Office of Research Ethics at the University of Toronto (Ref: 42337) and the Institutional Review Board of the Association for Social Development, Pakistan (Ref: IORG0011016 dated 04 March 2022).

### Setting

We will implement the study in rural Punjab, Pakistan. The Province of Punjab, Pakistan, has 110 million inhabitants in 36 districts. About two-third of the population live in rural areas and experience challenges in accessing health care such as physical distance, affordability, and social constraints for woman travellers. In many situations, access challenges are further complicated by a low quality of care in NCD diagnosis and treatment (29). In Punjab, the rural population is served mainly by a network of public health and primary care facilities consisting of more than 315 Rural Health Centres (RHC) and 120 District Hospitals. Each RHC serves a catchment population of approximately 150,000 – 250,000 rural inhabitants. These RHCs offer NCD diagnostic and treatment services, emergency obstetric and neonatal care, and treatment of general ailments such as malnutrition and arthritis. The RHC staff include at least: 1) three or more doctors; 2) a range of allied health workers including drug-dispensers, lady health visitors, nurses, and laboratory technicians; and c) support staff such as record clerks, store-keepers, office support staff, and cleaners. The Association for Social Development (ASD), a non-government organisation (NGO) and the local project lead, is leading a project funded by the World Diabetes Foundation (WDF) which has introduced diagnosis, treatment, and follow-up care for diabetes and hypertension in 84 RHCs in Punjab. RHCs provide outpatient services from 8am to 2pm during weekdays with emergency services available 24 hours a day. Patients who visit RHCs are mostly poor and prefer to use public health service because it is free of charge. Often, they do not use private consultations.

### Randomization and blinding

We will recruit and randomise 30 RHCs in three districts: Sarghoda, Jhang and Muzaffargarh, in rural Punjab. All three districts are a part of the WDF project which has already provided diagnosis and treatment care for hypertension and diabetes. RHCs will be stratified by district and further balanced on population using covariate constrained randomisation, and allocated into intervention and control at 1:1 ratio (30). With covariate constrained randomisation, many possible allocation plans are constructed. For each plan a balance score is calculated that represents the degree of balance on the covariates of interest. The final randomisation scheme is then randomly selected from the collection of plans with acceptable balance. In this case, it is selected from the top 500 plans with least imbalance on population. RHCs will not be blinded due to the nature of the intervention. To mitigate the risk of bias, we will: 1) mask the statistician team when data is presented and analysed; and 2) employ objective outcomes.

### Eligibility and recruitment

Eligible participants are patients who 1) are aged 25 years or older; 2) reside in the catchment of selected RHCs, 3) provide informed consent, 4) are newly diagnosed of hypertension, i.e., having a baseline blood pressure reading (recorded from the second blood pressure reading using a validated electronic blood pressure machine) of more than 140/90 mmHg; or who is an existing hypertensive patient but with uncontrolled blood pressure with a baseline blood pressure over 140/90 mmHg, and 5) have a smartphone or can access to a smartphone from a relative. Exclusion criteria include having an acute cardiovascular event in the last three months, terminal disease, or other conditions that the RHC staff determine impossible to participate. We will recruit patients into the studies via allied health workers who will screen patients for eligibility during their visit to RHCs, explain the study purpose, and obtain oral informed consent. Based on the NCD patient data reported from the RHCs, on average, 25-30 hypertensive patients are enrolled at a single RHC each month, we estimate that the recruitment will take two months when recruiting both new and existing patients. In total 1500 patients will be enrolled in the study. Sample size calculations are explained below.

### Procedures

#### Control arm

Our control arm will implement usual care, which is the routine hypertension and diabetes diagnosis and treatment under the WDF project. The WDF project provides initial Zoom-based training of NCD care to RHC staff, but no tele-mentorship is offered. Currently, NCD care is delivered through outpatient consultations where patients must visit RHCs to get screened, diagnosed, registered, and treated. RHCs stopped service from April to June 2020 due to the pandemic, but since August 2020, regular visits to RHCs have been largely resumed. Basic anti-hypertensive and anti-diabetic medicines including insulins are provided free of charge in RHCs. In case of shortage, patients are advised to come back or purchase from private pharmacies. Patients are referred to district hospitals for cardiovascular disease or COVID-19 diagnosis if suspected. Under the usual care, patients with hypertension or diabetes are required to visit RHCs every month to renew their medications and measure their blood pressure. No other interventional components will be implemented in the control arm.

#### Intervention arm

We will implement a comprehensive package of DHIs to connect patients, patient champions and public health providers to improve management of NCDs during the pandemic, including 1) providing training to health providers regarding an integrated NCD-COVID guideline; 2) using a smartphone app to improve NCD case management and linking with patient champions; and 3) employing tele-mentoring platform to improve quality of care (Fig 1). Patient champions are experienced patients who can provide peer support, which has been used by ASD in its tuberculosis project. DHIs in the above three categories are described in full below and then summarised according to the WHO classification of DHI categories as 1.0 (client/ patient) and 2.0 (health providers) for cross-reference (31).

**Fig 1.**
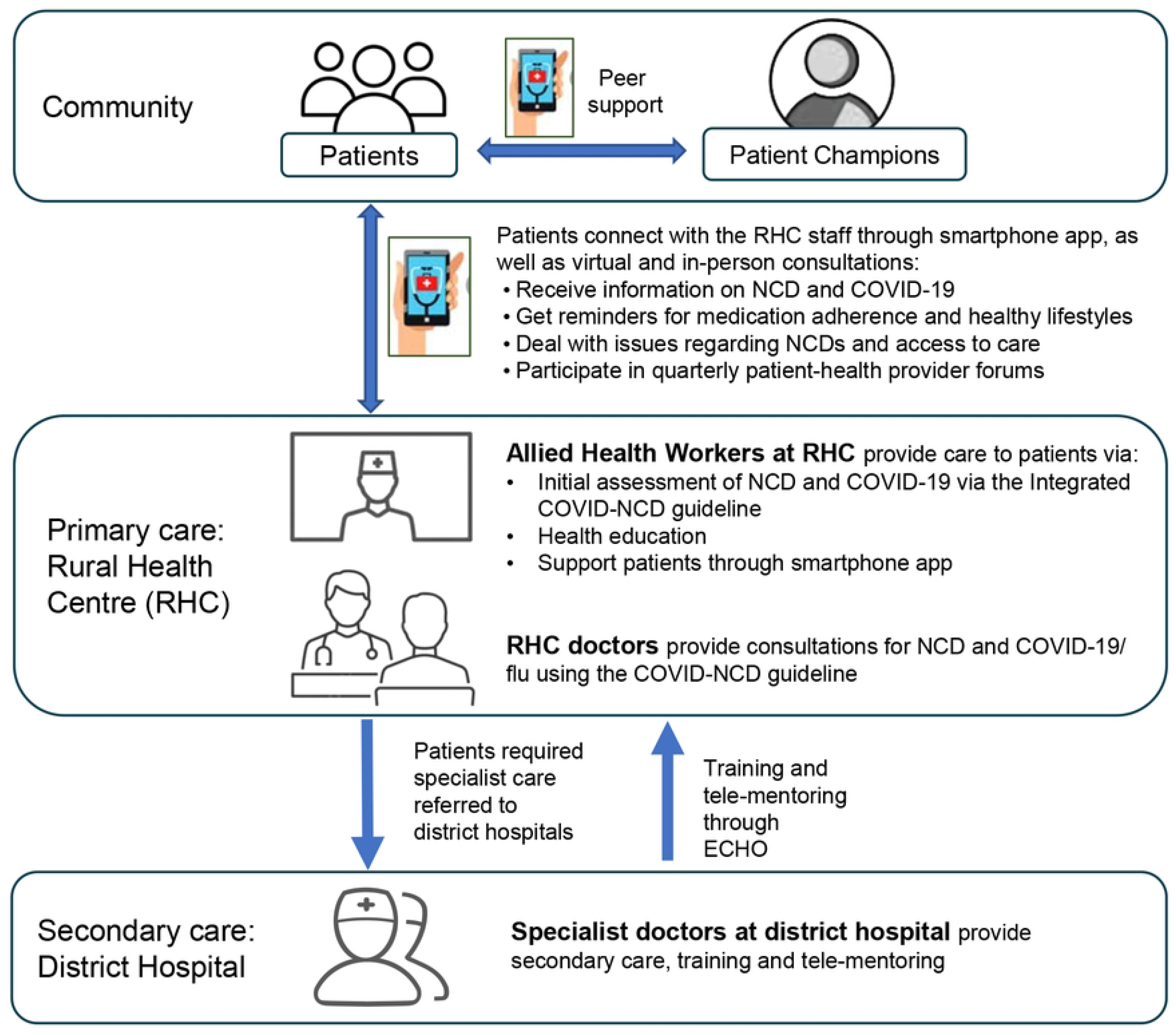
Diagram of intervention activities

#### Integrated COVID-NCD guideline

We have developed an integrated COVID-NCD guideline to support RHC staff to deliver care to patients with flu-like symptoms (for symptomatic SARS-COVID-2 and other influenza viruses) in terms of promoting rational use of personal protection equipment (PPE), promoting COVID-19 vaccination, and providing case management, health education, and home quarantine support. This guide is based on our previous experience in developing COVID-19 guidelines for primary healthcare facilities in Asia and Africa (32,33). The guideline integrates with existing hypertension and diabetes management guideline in Punjab. We also developed two versions of guidelines and training modules, one for the doctor and one for the allied health workers (WHO DHI 2.8) in RHCs. The training materials for allied health workers focus on using a smartphone app to improve patient retention to care, adherence to medications and healthy lifestyles such as reducing sugar intake, physical activity, and smoking cessation in local context (WHO DHI 2.2 and 2.4).

#### A smartphone app

We developed a smartphone app to link patients with RHC staff and patient champions. The app is an adaptation of OneImpact currently employed to provide management for patients with drug resistant tuberculosis in Pakistan. Patients will be provided the option of visiting RHCs every two months with prolonged prescriptions. Patients will be able to use the app to a) access knowledge on NCD and COVID-19/ flu care, including medication adherence and healthy lifestyle for managing hypertension and diabetes at home, and knowledge to prevent COVID-19 transmission, home management, infection control and when to seek health advice (WHO DHI 1.1); b) report any medical issues to allied health workers in RHCs (WHO DHI 1.5); c) receive virtual care (i.e., reminders and consultation calls) from allied health workers in RHCs based on their medical issues (WHO DHI 1.1, 2.2 and 2.4); d) if not responding to reminders nor reporting any medical issues, patient will receive follow-up calls from allied health workers during the gap month (WHO DHI 1.1, 2.2 and 2.4); e) participate in quarterly patient-health provider forums (WHO DHI 1.6); and f) participate monthly in peer-support sessions with patient champions for practicing recommended healthy behaviours, and addressing social issues in adherence to treatment and access to hospital and stigma (WHO DHI 1.3).

#### Tele-mentoring

We will conduct tele-mentoring provided by specialists in District Hospitals, based on the Extension for Community Healthcare Outcomes (ECHO), a remote training and mentoring system with proved effective to improve NCD management (34). We will employ the ECHO case-based learning approach using best practice and data within RHCs regarding NCD management, COVID-19 screening, prevention, and home management (WHO DHI 2.5 and 3.1).

Upon recruitment, patients in the intervention arm will be facilitated by an allied health worker on site regarding how to install and use the app. For patients who do not have access to a smart phone, or who have low smartphone literacy, one of their family members who has a smartphone or good smartphone literacy will be trained either on site or within a week of enrolment. We will identify “patient champions” who are persons with lived experience of well managed diabetes or hypertension in the catchment area and are willing to provide consultation support. We will have one male and one female patient champion in each district. Creation of study materials and intervention components will be grounded in a gender and culturally responsive approach, informed by local stakeholders and best practices, to support equitable delivery of services and workforce capacity building.

### Outcomes

The primary outcome is systolic blood pressure at 10 months measured in the RHC. Systolic blood pressure has been commonly used to measure hypertension management outcomes(35). It is also an important indicator in managing patients with diabetes (36,37). High blood pressure alone is a significant risk for cardiovascular disease(38). Our secondary outcomes include NCD related indictors such as: 1) diastolic blood pressure (mmHg) measured in the RHC at 10 months; 2) percentage of patients with controlled blood pressure measured below 140/90 mmHg for patients with hypertension but without diabetes, and below 130/80 mmHg for patients with diabetes at 10 months; 3) random blood glucose at 10 months (mmol/L); 4) body mass index (BMI) at 10 months; 5) number of consultations with RHC doctors, including both in-person and virtual consultations; 6) systolic and diastolic blood pressure measured at 6 months. We will also measure indicators related with COVID-19 including: 7) proportion of patients who have had at least three doses of COVID vaccinations; 8) proportion of participants been admitted to district hospitals for any reasons; 9) all-cause mortality rate during the 10 months. We are not able to measure better recognisable indicators for diabetes management such as haemoglobin A1c because the test is not available in RHCs and is costly.

#### Sample size

Sample size is based on demonstrating the intervention improves systolic blood pressure. A difference of 3 mmHg in systolic blood pressure at the population level is deemed to be the minimal clinically important difference to detect. Due to the pragmatic nature of this trial, which tends to attenuate treatment effects, a difference of 3 mmHg in this setting indicates an effective treatment (39). Based on prior work the standard deviation of systolic blood pressure is expected to be about 11 mmHg (40,41). We will further assume a modest intra-class correlation of 0.03 and set a Type I error probability of 0.05 and plan on 40 patients per cluster for the sample size estimation. Sample size calculations for the comparison of means in cluster trials indicate that 524 patients per group would be required for 85% power and 614 patients per group would be required for 90% power. Although 614 is slightly higher than the 600 expected from 15 clusters of 40 each, the adjustment of baseline systolic blood pressure in the analysis will increase the power relative to the simple unadjusted analysis reflected in the sample size estimation. Considering a 20% loss-to-follow-up rate based on our previous trials in Pakistan and China we aim to increase to 50 patients per cluster, that is a total of 1500 patients to be recruited (23,42). We do not anticipate loss-to-follow-up at the cluster level. Power will increase if all clusters meet the enrolment target of 50 patients.

### Internal pilot process

We will assess the feasibility of implementing the intervention and running our trial processes during an internal pilot study that will be conducted after the randomisation but before the main trial. The pilot intervention will be conducted in one RHC of the intervention arm and one RHC in the control arm out of the three selected districts. The internal pilot study aims to assess recruitment rates and the extent to which the intervention is delivered within RHCs; it will also contribute outcome data to the main trial. In the intervention RHC, we will pilot train RHC staff using the integrated COVID-NCD guideline and demonstrate how to use the smartphone app for consultation and communication with patients. We will recruitment patients over the course of one week to test feasibility of recruitment. We will then follow up patients for two weeks to understand if patients are able to use the app for health education and communication with health providers. We will also train patient champions and conduct a peer support session to test its feasibility. All intervention and data collection tools will be pilot tested and revised based on feedback. In the control RHC, we will pilot the data collection tools, recruit patients over a week and follow up for two weeks to understand recruitment and retention. The two RHCs in the pilot study will be recruited and followed-up for one month, and the decision as to whether to continue with the full trial will then be decided based on two key criteria: 1) sufficient levels of recruitment; 2) acceptability and feasibility of interventions (at least 50% of health providers trained and 50% using the smartphone app and integrated COVID-NCD guidelines at the end of the month). If these criteria are met, the internal pilot RHCs and their outcome data will then become part of the main trial,

### Process evaluation

Using the RE-AIM framework, we aim to describe the health system and service delivery context in which the intervention is implemented, explore delivery and dose of intervention components received, and understand mechanisms of impact and unanticipated consequences on health providers and parents (43). Methods will include document review (e.g., meeting minutes, programme notes, record reviews etc), training session observations, and qualitative interviews with stakeholders. Specifically, we aim to understand: 1) Reach: the recruitment process and programme inclusion; 2) Effectiveness (feasibility and acceptability): to what extent our measures are understood and accepted by patients; 3) Adaptation: changes of interventions made at the facility level; how the NCD-COVID care connects with other programmes such as tuberculosis and immunisation; and any resistance from structural, professional or personal factors in adopting the change; 4) Implementation: issues related to fidelity/quality of implementing the NCD-COVID programme, and any unanticipated consequences; 5) Maintenance: to what extent the NCD-COVID programme is institutionalized in different RHCs. Our previous similar work will inform this evaluation (44–46). Detailed methods for sampling, data collection and analysis will be produced later following local consultations with the intention of balancing methodological rigor with an approach responsive to gender and equity. The data will be analysed using a thematic approach reported based on Consolidated Criteria for Reporting Qualitative Studies (COREQ) (47,48).

### Cost-effective analysis

An incremental cost effectiveness analysis will be conducted to estimate the added benefits of the intervention regarding extra reduction of 1 mmHg patient systolic blood pressure in the intervention arm compared with that in the usual care from a societal perspective, which entails an assessment of all resource costs irrespective of payer. Costs associated with intervention over and above usual care (excluding research costs) will be collected and used together with the above effectiveness/outcome measures to calculate the incremental cost-effectiveness ratio(48). Costs will include capital costs such as training, other costs to strengthen the capacity of providers to deliver enhanced services, and recurrent costs such as drugs and laboratory tests. Salaries of staff in RHCs will be included in case of extra working time is sought in some clusters during the intervention. Project logbook review and interviews with managers will be conducted to obtain cost data. Hospitalisation and consultation costs for any medical reasons during the trial period, including travel and waiting time, of patients and their relatives will be recorded and converted into monetary forms using conventional age-sex adjusted market wages. We will use net benefit regression framework which will allow us to adjust for potential confounders and to calculate the clustered standard errors using the sandwich variance estimators to account for a clustered randomised design (49). Markov models will be used to simulate possible future states of the intervention, and to capture possible longer-term health outcomes (e.g., additional years of life) and associated costs of care (50).

#### Data collection

We will measure patient blood pressures, random blood glucose, and patient body weight and height at randomisation and during each in-person consultation at RHC. We will remind patient to visit RHCs for the last measure at 10 months. During each consultation, the RHC doctor will measure participants’ blood pressure using a calibrated and standardised electronic sphygmomanometer after five minutes of seated rest for two times with an interval of two minutes. If the difference in the two consecutive readings is over 5 mmHg, a third measure will be taken. The lower readings in systolic blood pressure of the last two measures, together with the diastolic blood pressure measured with the determining systolic blood pressure, will be recorded as patient blood pressure. Doctors will also measure body weight and height using standardised scales. Blood samples will be collected and analysed in RHC according to national guidelines for random plasma glucose. We will record all biomedical indicators on a revised treatment card that is routinely used in RHCs, including patient basic demographics, smoking status, COVID-19 and NCD diagnosis (including hypertension, diabetes, chronic obstructive pulmonary disease, cancer, and cardiovascular disease), consultation dates (including both physical and virtual), and medication prescriptions. We will conduct a questionnaire survey at 10 months by trained ASD staff to collect patient socioeconomic status, disability condition, and costing data related with medical consultations, medicine, laboratory tests and any hospitalisation, as well as any opportunity cost. Patients will be approached in the community if not presented in RHCs after three reminders or loss-to-follow-up during the trial, and post-posthumously with their close family members for patients who died during the trial. COVID-19 vaccination information will be collected from the National Vaccination Database.

#### Statistical analysis

We will present appropriate descriptive statistics for RHCs and at the individual level, along with appropriate summary statistics and their associated 95% confidence intervals for all the outcomes. Then, for all outcomes, we will produce a main set of estimates of intervention effectiveness using cluster-level methods of analysis suitable for cluster trials. In particular, a linear mixed effect model will be used to analyse the primary outcome (systolic blood pressure) adjusting for the baseline systolic blood pressure, district (the stratification factor), and patient level factors (socioeconomic status, religious and disability). For other continuous secondary outcomes a similar analysis will be employed. Binary secondary outcomes will be analysed with the generalised estimating equation (GEE) approach, employing a binomial family with logit link function and exchangeable working correlation structure on sites. There will be no interim analyses. Intent to treat analysis will be conducted. An inverse probability weighted analysis will be conducted in the event of significant loss to follow-up (more than 10%). A formal statistical analysis plan will further describe the analysis, including any other pre-specified covariates to adjust for.

#### Analysis of process evaluation

Both quantitative and qualitative data will be analysed. The implementation data from the trial, including program logbooks and app usability data, will be summarised, described and analysed using appropriate statistical methods. The qualitative data will be collected using a descriptive approach where information is gathered by interviewing those experiencing the phenomenon under investigation as part of a mixed methods approach (51). Notes will be reviewed after each interview to identify emergent topics and allow for exploration in subsequent interviews. Data will be translated and transcribed, then analysed using NVivo 10. Qualitative data analysis will use a thematic approach to group information from the document review and stakeholder interviews into themes guided by constructs in the RE-AIM framework.

#### Trial management

A data management committee will be established to 1) safeguard the safety and privacy of patients involved, and to 2) ensure that all data are collected according to agreed ethical guidelines, properly stored and only used for research purposes. A trial management unit (TMU) will be established in Islamabad, Pakistan with a trial manager and one staff locally and a research coordinator from University of Toronto. TMU will manage the day-to-day activities of the trial. Prof Xiaolin Wei from the University of Toronto and Dr Muhammad Amir Khan from ASD will be the co-guarantors of the trial who have full access to the trial dataset. Prof. Kevin Thorpe will be the lead statistician. A trial steering committee will be established and led by external members. We will organise virtual trial steering committee meetings at the beginning of the study, and then every six months. The committees may also meet on an ad hoc basis should needs arise.

## Discussion

To our knowledge, this trial is one of the first to evaluate the effectiveness of a comprehensive digital health intervention package in improving NCD management and health system resilience in LMICs. Ambitious action to tackle NCDs is crucial given that most LMICs are falling short of the targets set out in Sustainable Development Goal 3.4 to reduce NCD mortality by a third between 2015 and 2030. The pandemic has further derailed progress, both through the disproportionate impact of COVID-19 on people with NCDs, as well as through the massive health systems and NCD care disruptions resulting from COVID-19 response efforts. Similar disruptions of NCD care were seen in the recent floods. Not only have these disruptions affected people already living with NCDs, but limited access to screening and diagnostics during the pandemic means many more are undiagnosed and presenting to care with higher acuity, complexity, and in need of comprehensive primary services.

Our interventions fit well into global calls, as well as local priorities in Punjab for implementing and scaling up cost-effective digital solutions for NCD care in primary care settings (8,52). Based on this, we developed the smartphone app based DHIs to improving patient-provider communication. The app was adapted from one that has already been employed for managing MDR-TB patients in Pakistan. Through the app, patients will receive integrated NCD-COVID care virtually, therefore reducing half of their routine in-person visits to RHCs while getting more rapid feedback from RHC staff. We will also provide patient peer support and patient provider communication on a regular basis. We will engage the NCD programme at the national, provincial and district levels in developing guidelines and training modules to ensure the materials are ready to be adapted to other remote areas and the project can be scaled up in Pakistan. In addition, the trial is designed as a hybrid trial that will test both the evidence, the comprehensive DHI package, and the implementation strategy, with an aim to facilitate future scale-up other LMICs. We will employ the RE-AIM framework in our process evaluation and a cost-effectiveness analysis to provide evidence on implementation, adaptation, maintenance and scalability of this DHI package in similar resource constraint settings.

The study has several limitations. First, it is an open label trial where both health providers and patients cannot be blinded, which will introduce biases on performance of both health providers and patients. However, we will minimise any of the bias by collecting objective patient indicators using standardised equipment purchased by the project. We will blank the allocations from the statisticians in data analysis. Second, there is a risk of non-compliance of health providers and patients. We will provide on-site supervision and the ECHO-based tele-mentoring with health providers to identify non-compliance and provide timely feedback. Patient champions will connect patients or their relatives who have low literacy in smartphones and who do not attend virtual consultations. In addition, we will provide gender sensitive support in matching the patient champions and patients. Third, Pakistan is currently under a national emergency of severe flooding. Though Punjab is largely not affected, it may cause shortage in staff and funding for NCDs. Our interventions will help the public health system to strengthen their resilience to respond to different disasters. We will document the context and any changes and continue to engage with the provincial NCD program for possible way of mitigations.

In conclusion, we present the protocol of a pragmatic cluster randomised controlled trial to evaluate the effectiveness of a comprehensive DHI package to improve management of NCD and improve resilience in public primary care facilities in rural Punjab, Pakistan. The trial is co-designed with Pakistani NGO and governmental stakeholders, in line with the WHO’s framework on integrated people centred health services to institutionalise DHIs to respond to health system challenges. If proved successful, we aim to adapt and scale up the DHI package in rural Pakistan and other LMICs with similar challenges.

## Trial status and timelines

The trial is registered at: ClinicalTrials.gov (Identifier: NCT05699369)

### Timelines

The study will be done over a 24-month period, with a nine-month preparation and pilot phase, a two-month patient recruitment, 10 months month treatment phase for all patients, and a three-month data analysis and write-up stage. We will disseminate trial results through research articles and policy briefs. See Fig 2 for details of the trial timeline.

**Fig 2.**
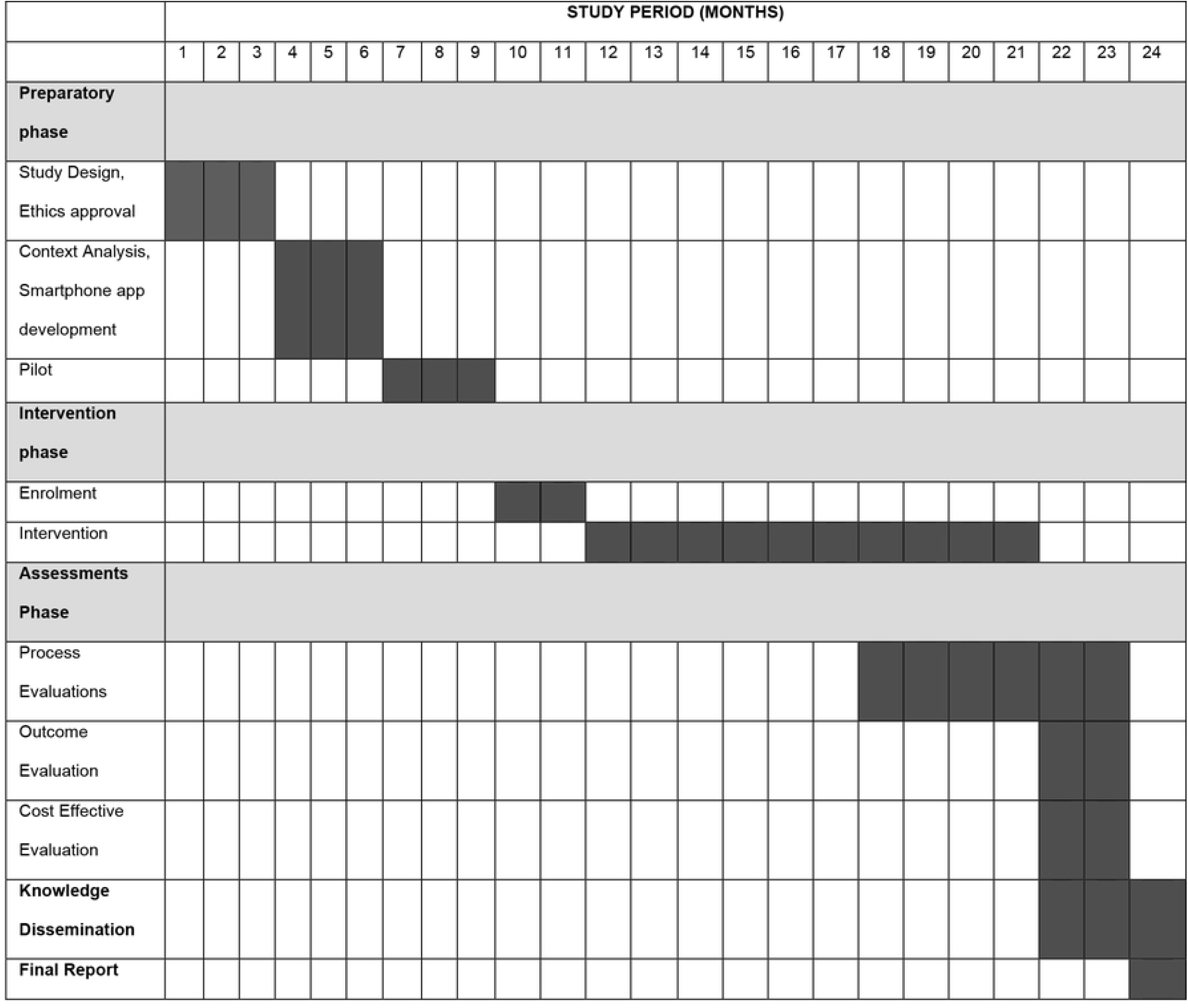
Trial timelines

## Data Availability

No datasets were generated or analysed during the current study. All relevant data from this study will be made available upon study completion.

## List of abbreviations

ASD: Association of Social Development
DHI: Digital Health Intervention
ECHO: Extension for Community Healthcare Outcomes
RHC: Rural Health Centre
NCD: Non-Communicable Diseases
LMICs: Low-and-Middle-Income Countries
TMU: Trial Management Unit
WDF: World Diabetes Foundation
WHO: World Health Organisation

## Contributors

All authors made substantive contributions to the trial development and provided final approval for this manuscript. XW, MAK, NK and JDW designed the trial and related studies. XW drafted the manuscript. NK, NM, HD contributed to the pilot study. KT contributed to the statistical issues in the study design. JW, NK, NM, HD, SG and GA contributed to trial intervention materials and critically reviewed the manuscript. XW, VH and EG contributed to ethics development. WD and VH contributed to the qualitative study. JW contributed to digital health intervention review. NK and AL contributed to the health economic evaluation. XW and MAK are co-principal investigators of the trial.

## Acknowledgement

We thank the Steering Committee Members: Mr. Arif Muneer (Pakistan and the Chair of the Committee), Dr. Haroon Jehangir (Former Directorate General of Health Punjab Pakistan), Dr. Shahzad Khan (Health Services Academy Pakistan), Dr. Muhammad Amir Khan (ASD, Pakistan) and Prof. Xiaolin Wei (University of Toronto). We thank Dr. Gary F. Lewis from Toronto General Hospital, and Dr. Bruce B. Strumiger from ECHO Institute, University of New Mexico for their support. We also acknowledge all the health providers, patients, and patient champions who participate in discussions to develop the proposal.

## Declarations

Ethical approvals and consent to participate: The trial protocol has obtained ethical approval from the Office of Research Ethics at the University of Toronto (Ref: 42337) on 23 March 2022and the Institutional Review Board of the Association for Social Development based in Pakistan (Ref: IORG0011016 dated 04 March 2022).

Informed consent forms with patients’ preference of language (Urdu or English) will be collected before any patient is recruited into the study. The healthcare providers will fully explain the information sheet and consent form to the participant in Urdu or English. Both the participant and the healthcare provider are required to sign the consent form. If the participant is not able to sign due to illiteracy, they can use their thumbprint instead. The healthcare provider will keep the signed informed consent form and leave a copy to the participants for their reference.

## Consent for publication

Not applicable.

## Availability of data and materials

Not applicable as the protocol doesn’t report any results.

